# Subarachnoid Hemorrhage Trials: Cutting, Sliding, or Keeping mRS Scores and WFNS Grades

**DOI:** 10.1101/2023.09.28.23296257

**Authors:** Akshitkumar M. Mistry, Jeffrey Saver, William Mack, Hooman Kamel, Jordan Elm, Jonathan Beall

## Abstract

Rigorous evidence generation with randomized controlled trials (RCTs) has lagged for aneurysmal subarachnoid hemorrhage (SAH) compared to other forms of acute stroke. Besides its lower incidence compared to other stroke subtypes, the presentation and outcome of SAH patients also differ. This must be considered and adjusted for in designing pivotal RCTs of SAH patients. Here, we show the effect of the unique expected distribution of the SAH severity at presentation (World Federation of Neurological Surgeons, WFNS, grade) on the outcome most used in pivotal stroke RCTs (modified Rankin Scale, mRS) and consequently on the sample size. Further, we discuss the advantages and disadvantages of different options to analyze the outcome and control the expected distribution of WFNS grades in addition to showing their effects on the sample size. Last, we offer methods that investigators can adapt to more precisely understand the effect of common mRS analysis methods and trial eligibility pertaining to the WFNS grade in designing their large-scale SAH RCTs.

**Purpose:** The generation of rigorous evidence to inform the management of patients with aneurysmal subarachnoid hemorrhage (SAH) has lagged other types of acute strokes. The reason for this lag is multifactorial—one being that SAH has the lowest incidence of all forms of stroke. However, the paucity of SAH randomized controlled trials (RCTs) can also be self-exacerbating. Rather than adopting existing trial designs and biostatistical methods, it forces new investigators to craft these anew. Here, we provide a basic biostatistical guide for investigators to navigate two foundational dilemmas in designing large-scale SAH RCTs.

## Measures of SAH Severity and Outcome

The severity of presentation universally predicts patients’ functional outcomes in stroke. The ordinal World Federation of Neurological Surgeons (WFNS) scale^1^ is one scale used to grade the SAH severity, increasing from 1 to 5. Although the Glasgow Outcome Scale has been used for SAH, the archetypal outcome used to measure neurological morbidity and mortality in stroke patients is the ordinal modified Rankin Scale (mRS), ranging from 0 (no neurological disability) to 6 (death). For a biostatistically-informed trial, it is critical to estimate the expected distribution of WFNS grades, mRS scores, and correlation between the two.

## Expected WFNS Distribution is Skewed

The expected distribution of WFNS grades is largely skewed toward grade 1. For exemplary purposes, we retrieved a few large RCTs with broad eligibility criteria and cohort studies from different regions of the world (which should possess less selection bias common to research studies) and estimated the proportion of WFNS grade 1 to 5 (in %): 41, 22, 7, 15, 15 (Figure 1A; Supplemental Figure 1), with grade 2 being the median. For each WFNS grade, we acquired the outcome on the full mRS scale from the ULTRA^2^ RCT (Figure 1B). We chose ULTRA because it is the most recent (2021) pivotal SAH RCT of 955 patients treated with the current standard of care. Collectively, the skewed expected WFNS distribution and the correlation between WFNS grade and mRS outcome in SAH patients may pose challenges. The effect of any intervention in WFNS grade 1 patients will be diluted because 75% of them will already have a good outcome (commonly accepted as mRS 0-2; Figure 1B). Conversely, unless an intervention has strong therapeutic effects sufficient to improve substantial brain injury in WFNS grade 5 patients, the effect size will also be diluted because 76% of them will have a bad outcome (mRS 4-6). Since interventions confirmed to show a therapeutic benefit for SAH patients in RCTs are scarce,^3^ the design of large-scale SAH RCTs must be optimized to detect a therapeutic effect considering variable effects based on WFNS grade.

**Figure 1.**
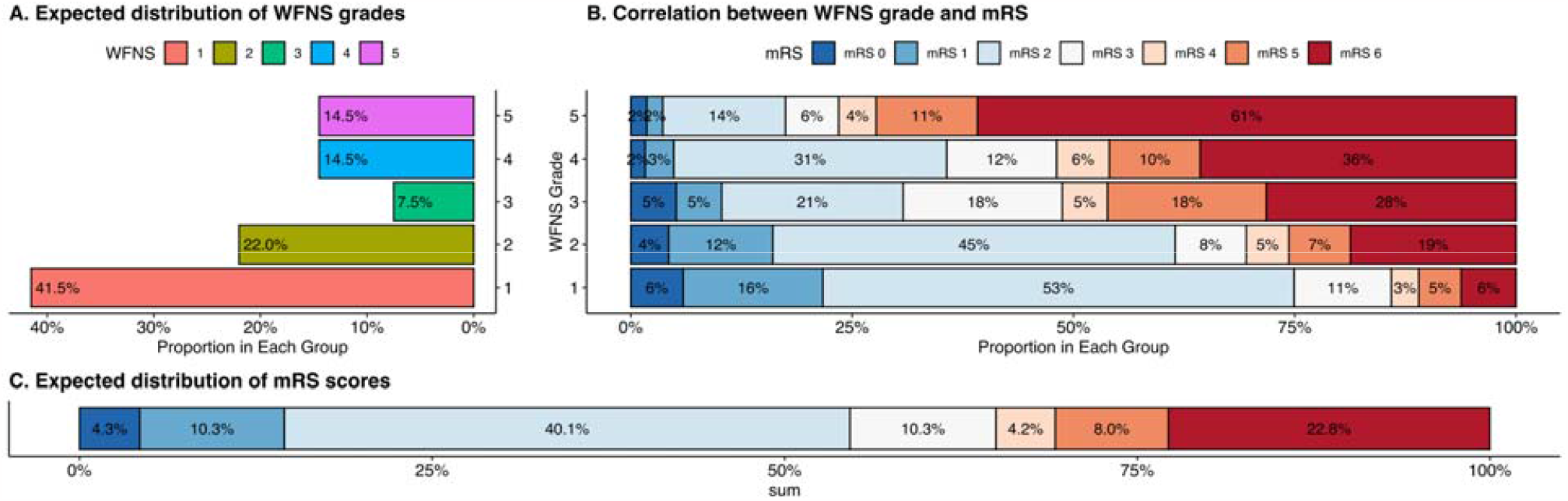
Expected distribution of World Federation of Neurological Surgeons (WFNS) grades (A) and modified Rankin Scale (mRS) scores for subarachnoid hemorrhage patients (B,C). Data in (A) are derived from a weighted average of 8 large studies in Supplemental Figure 1 and (B) from the ULTRA trial.

## Outcome: Cut, Slide, or Keep mRS Scores?

Deciding how to analyze the mRS scale is a frequent dilemma. Common options are 1) fixed dichotomization, focusing on either good outcomes (mRS 0-2 vs. mRS 3-6) or bad outcomes (mRS 4-6 vs. 0-3); 2) “responder” analysis with a sliding dichotomy, prespecifying what constitutes a good outcome depending on the presenting SAH severity (for example, mRS 0-1 for WFNS grade 1, mRS 0-2 for grade 2-4, and mRS 0-4 for grade 5); and 3) “shift analysis” across all mRS scores analyzed with an ordinal regression.

We studied the impact of these options on the sample size for a two-group pivotal SAH trial (80% power; two-tailed α 0.05) using the baseline (control) mRS distribution dictated by expected WFNS distribution (Figure 1B-C). While fixed dichotomy at mRS 0-2 and mRS 4-6 is widely accepted, there are no precedents or acceptable, validated schemes based on WFNS grades for sliding dichotomy. Therefore, we generated all reasonable permutations of sliding dichotomy schemes by limiting a good outcome of WFNS grade 1 to mRS ≤2 and grade 5 to mRS 3-4. An mRS 0-2 for WFNS grade 5 is not only ambitious but also numerically constricts the “slide” and thus greatly limits the sliding options because the fundamental principle of a sliding dichotomy is cutting (or dichotomizing) the mRS scale at the same or higher mRS score for sequentially higher WFNS grades. For example, allowing dichotomization of WFNS grade 5 at mRS 2 will limit WFNS grades 1-4 to be dichotomized at mRS ≤2 only (i.e., a “quicker” slide) as opposed to if WFNS grade 5 is dichotomized at mRS 4, then WFNS grades 1-4 can be dichotomized broadly along mRS scores ≤4 (see examples in Supplemental Figure 2). With these rules, 99 sliding dichotomy permutations or schemes were generated. Thus, including the two commonly used fixed dichotomy schemes, we tested a total of 101 dichotomy schemes.

For exemplary purposes, we considered the common effect size of 10% absolute change in the dichotomized outcomes. (For reference, Supplemental Figure 3 reviews the difference between absolute and relative effect sizes.) Using the expected mRS distribution from Figure 1C as the control group, a fixed dichotomy at mRS 0-2 requires 14.6% more patients than at mRS 4-6 (754 vs. 658; see Figure 2A blue versus purple lines at 10% effect size). Sliding dichotomy schemes generally lower the required sample size compared to fixed dichotomy for a 10% absolute effect size. The sample size required with 49 of the 99 sliding dichotomy schemes is less than that required for mRS 4-6 fixed dichotomy, and 11 sliding dichotomy schemes require a sample size greater than that required for mRS 0-2 fixed dichotomy. Their sample sizes depend on how quickly the “slide” occurs from WFNS grade 1 down to 5 (blue dots in Supplemental Figure 4). The lowest sample size (340) is for a scheme setting a good outcome for grades 1-4 at mRS 0 and grade 5 at mRS ≤3, denoted as (0,0,0,0,3) for each sequential WFNS grade, while the highest (778) is for a scheme setting a good outcome for grade 1 at mRS ≤1 and grades 2-5 at mRS ≤4, denoted as (1,4,4,4,4). Last, if one intends to use all the information across the full mRS scale rather than dichotomizing it, then the sample size can only be estimated if the control mRS distribution and the expected mRS distribution for the intervention are known. The sample size may be greater or lesser than the dichotomy schemes, being entirely dependent on the two distributions, and the latter is often unknown. If a constant therapeutic effect is expected across all mRS scores (the practical validity of this assumption is highly arguable), then one may generate an expected mRS distribution for the intervention using a constant log odds shift (i.e., effect size; Figure 2B). With this assumption, a constant log odds shift generally yields a lower sample size than fixed dichotomies (Figure 2C).

**Figure 2.**
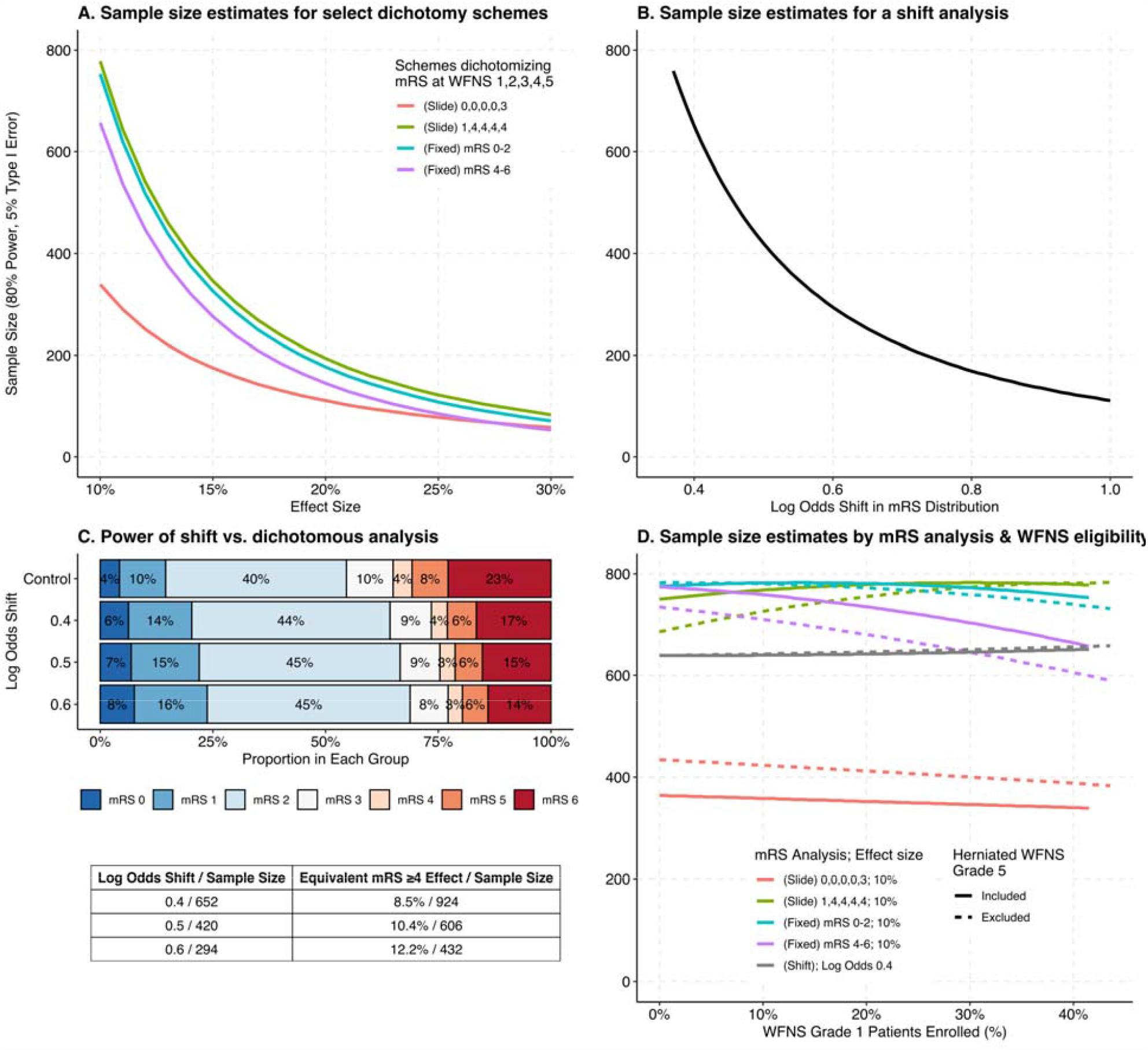
Samples size calculations for a two-group pivotal subarachnoid hemorrhage trial. WFNS=World Federation of Neurological Surgeons; mRS=modified Rankin Scale. Sample size estimates for (A) two examples of sliding and two fixed dichotomized outcomes using a two-sample test of proportions and (B) shift analysis using the Wilcoxon-Mann-Whitney test and control distribution in Figure 1C and (C). The sliding notation, for example, 1,4,4,4,4 designates the dichotomizing mRS score (≤) for each WFNS grade sequentially. (C) compares sample sizes of shift analyses by equating them to the effect sizes in a dichotomous analysis of mRS. (D) Sample size estimates based on WFNS grade 1 enrollment and grade 5 eligibility.

We focused on the influence of these outcome constructions on the sample size because sample size is concrete and often drives resources and cost. However, the decision should ultimately be multifactorial. The advantages and disadvantages of these approaches have been discussed in this reference.^4^ For the unique expected control distribution of mRS scores for SAH, it is critical to consider where is the greatest numerical and/or theoretical potential to detect a therapeutic effect based on the biological effect of therapy (i.e., further improving the outcomes of the large portion of mRS 2 patients, or decreasing mortality, the second largest portion of patients—or both; Figure 1C). Last, utility-weighted mRS is an option, but it is not well-studied or measured in aSAH patients,^5^ who receive different medical and surgical treatments compared to ischemic stroke patients.

## Eligibility: Cut, Slide, or Keep WFNS Grades?

Eligibility criteria can influence a trial’s sensitivity to detect a therapeutic effect. As discussed above, interventions could be less therapeutic on either end of the WFNS grade spectrum, thus including all WFNS grades may produce a diluted effect size of the intervention. While outright excluding WFNS grade 1 and/or 5 patients in pivotal SAH RCTs (which make up ∼55% of SAH patients) is an option, we caution against it to maintain the generalizability of the intervention given the lack of therapies with proven benefits in SAH patients and to not prolong the study with a lower the enrollment rate. A compromising option is to limit the enrollment of patients on either end of the WFNS grade spectrum. Although this reduces the enrollment rate and may dilute the treatment effect, it maintains some level of generalizability. Given that ∼40% of the SAH patients are WFNS grade 1, their enrollment could be limited to a percent that is deemed substantial for applicability of the results in clinical practice that is deemed to not drastically slow enrollment. Similarly, WFNS grade 5 patients can be limited, for example, using a different approach. Recent multicenter studies have shown that WFNS grade 5 patients can be further prognostically subclassified using the ‘herniated WFNS’ scale.^6^ Herniated WFNS grade 5 make up ∼30% of the WFNS grade 5 patients, and an overwhelming 88% of them have a poor outcome (mRS 4-6). The primary disadvantage of limiting WFNS grade 5 patients to only non-herniated WFNS grade 5 is the uncaptured applicability and missed opportunity to show the benefit of the intervention in this subpopulation with high mortality. We evaluated the effect on the sample size of limiting WFNS grade 1 from ∼40% to 0 and excluding herniated WFNS grade 5, comparing them to the default option of keeping all patients. We estimated the sample size of a two-group pivotal trial testing a 10% absolute effect size (80% power; two-tailed α 0.05).

Counterintuitively, in most cases, limiting WFNS grade 1 and excluding herniated WFNS grade 5 patients each increases the sample size compared to keeping all patients (Figure 2D). Limiting WFNS grade 1 decreases the sample size (max by -56) across the spectrum down to 0% in only 6 of the total 101 fixed or sliding dichotomy schemes. The maximum increase is 126. Excluding herniated WFNS grade 5 patients does not change the previously established dependence of sample size of the sliding dichotomy schemes on how quickly the “slide” occurs from WFNS grade 1 down to 5 (Supplemental Figure 4). The “quicker” and “slower” sliding dichotomy schemes experience a greater change in sample size. The largest increase in sample size (70) by excluding herniated WFNS grade 5 patients occurs in a scheme setting a good outcome as mRS 0 for grades 1-4 and mRS ≤3 for grade 5 (0,0,0,0,3), while the largest decreases occur in schemes setting mRS ≤1 for grade 1 and mRS ≤4 for grades 2-5 (1,4,4,4,4; -64) and the fixed dichotomy at mRS 0-3 vs. mRS 4-6 (-68). Excluding herniated WFNS grade 5 patients in 30 of the 101 dichotomy schemes decreases the sample size across the range of WFNS grade 1 proportions. The combined effect of excluding both WFNS grade 1 and herniated WFNS grade 5 is an increase in the sample size (max +172) in 87 schemes, including the fixed dichotomies, and a decrease (max -92) in 14 schemes. Of the fixed dichotomies, mRS 0-2 vs. mRS 3-6 is less sensitive to any of these single or combination of these approaches (min 732, max 784). Last, the sample size for a test (Wilcoxon-Mann-Whitney or proportional odds regression) utilizing all the information across the full mRS scale is relatively independent of the two WFNS eligibility approaches we discuss simply because the sample size is entirely dependent on the control and expected distributions. The latter often is unknown. Even predicting the expected distribution with a constant log odds shift in the control distribution will have minimal effect on the sample size.

Overall, these results seem counterintuitive. By limiting WFNS grade 1 and 5 patients, one may expect a lower sample size with a greater effect size of the intervention. However, we did not re-estimate the sample size with a higher expected effect size, which would decrease the sample size, because often preliminary estimates of the latter are unavailable. Second, these are purely mathematical results. The sample size needed to test an absolute 10% decrease (i.e., effect size) with the two-sample test for proportions is greatest when the control proportion is 55% (Supplemental Figure 5). Because of the skewed WFNS distribution and its dependent control mRS distribution, limiting WFNS grade 1 and 5 patients results in the control proportion approaching 55% in most dichotomy schemes. If the effect size of the intervention being tested has not been estimated with a preliminary study limiting WFNS grade 1 and 5 patients but is expected to be higher, we suggest an interim analysis that best fits the study to save resources.

## Conclusion

The expected distribution of WFNS scores skewed to the lowest grade dictates the sample size based on the primary analysis approach. There are advantages and disadvantages of different analysis approaches (fixed or sliding dichotomy and using the full mRS scale) and options to modify the expected WFNS distribution in a trial. Greater sample sizes are required for fixed dichotomies (especially mRS 0-2 vs. 3-6) and while most of the sliding dichotomies yield a lower sample size, there is no established precedent for picking one scheme over the other. Although sample size estimates are more precise when using all information across the entire mRS scale, this requires that both the control and expected distributions are known. While estimating the latter with a constant therapeutic effect across all mRS scores can be done and offers the greatest power compared to dichotomous schemes, the practical validity of this approach is arguable. Generally, limiting WFNS grade 1 enrollment and excluding justified subpopulations of WFNS grade 5 to increase the effect size of the intervention being tested will also increase the sample size. We provide the R code (Supplemental Content) that investigators can adapt to their expected WFNS and mRS distributions to navigate the two foundational dilemmas more precisely in designing large-scale SAH RCTs.

## Data Availability

All data produced in the present study are available upon reasonable request to the authors.

## Acknowledgments

We thank René Post, MD, PhD, Department of Neurosurgery, Neurosurgical Centre Amsterdam, Amsterdam University Medical Centres, Amsterdam, the Netherlands, for providing aggregated data from ULTRA.

## Sources of funding

NINDS U01NS087748 (J.E. and J.B.)

## Disclosures

None

## SUPPLEMENTAL CONTENT

**Supplemental Figure 1.**
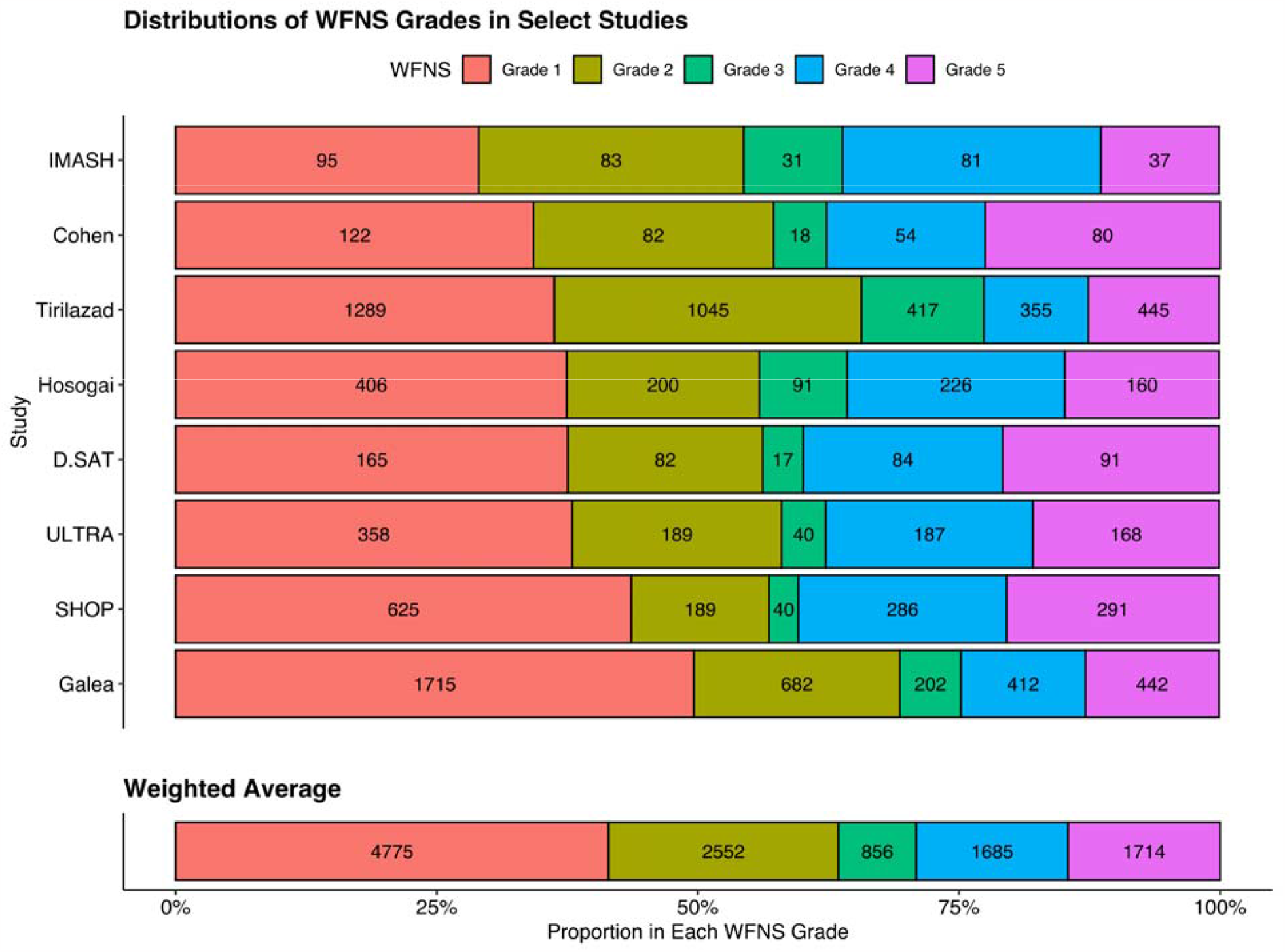
Estimation of expected World Federation of Neurological Surgeons (WFNS) grade in SAH patients. WFNS grade distributions from selected 8 studies are depicted. The weighted average is calculated and also plotted in manuscript Figure 1A. The references for these studies are listed below.

---

**Supplemental Figure 2.**
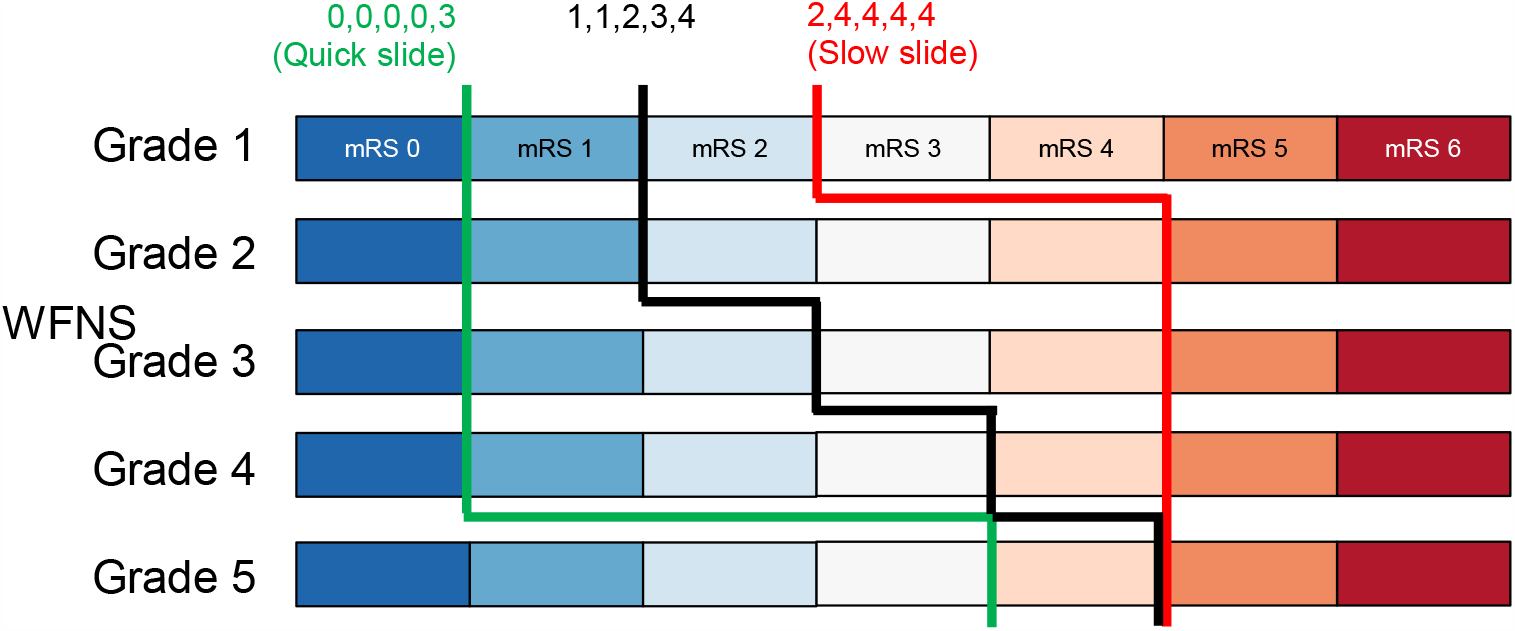
Examples of sliding dichotomies. Showing in green and red are quick and slow slides. The notation, for example, 1,1,2,3,4 designates the dichotomizing mRS score (≤) for each WFNS grade sequentially.

**Supplemental Figure 3.**
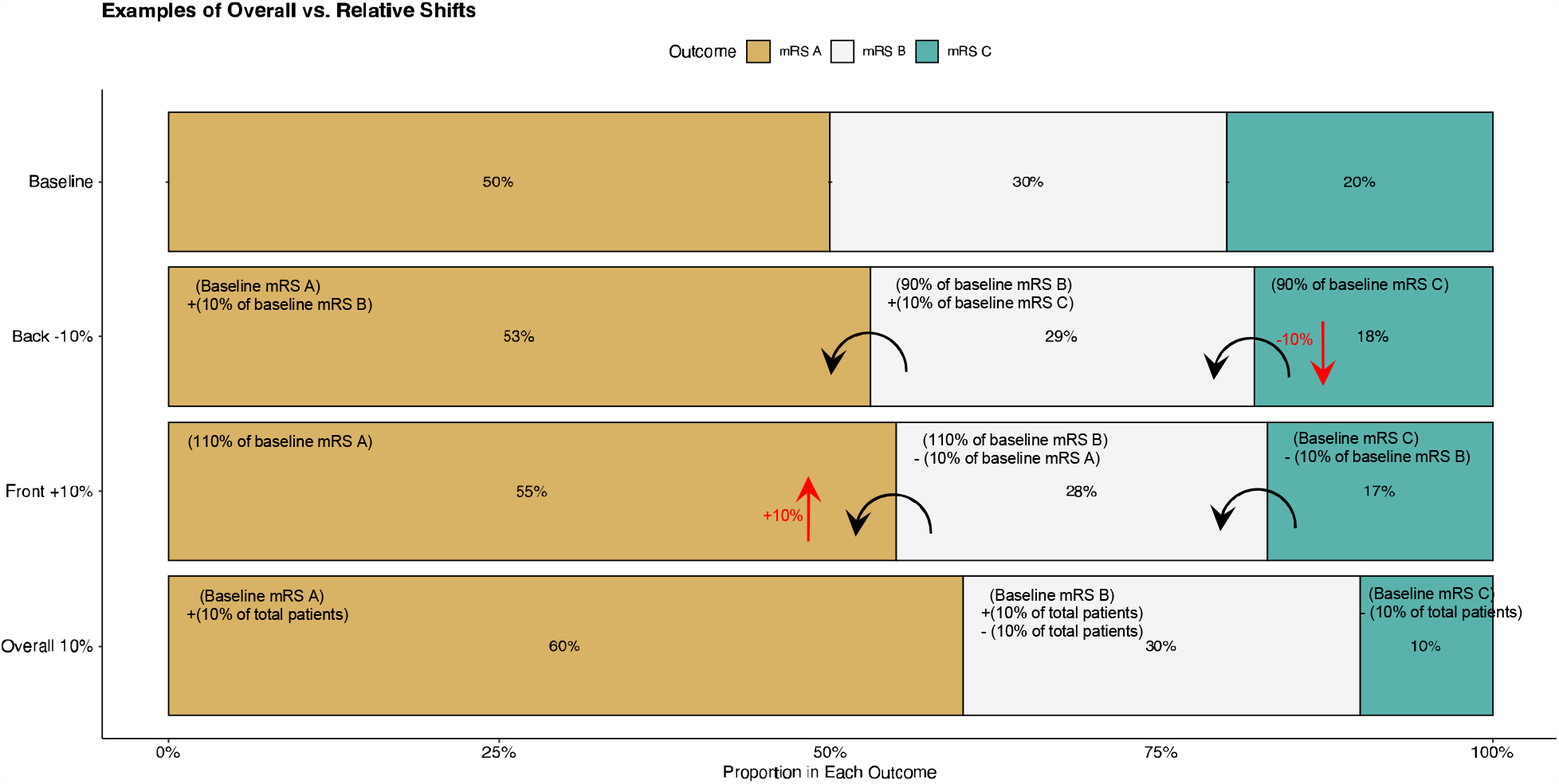
Examples of overall (absolute) vs. relative shifts. For exemplary purposes, we crafted a 3-scale ordinal mRS outcome (A, B, C). The control (baseline) distribution is shown with 50% of the patients having outcome A, 30% B, and 20% C. “Back - 10%” depicts the resulting distribution from the baseline by starting to decrease the proportion of patients with outcome C by 10%. “Front +10%” depicts the resulting distribution from the baseline by starting to increase the proportion of patients with outcome A by 10%. “Overall 10%” depicts the resulting distribution of a 10% absolute shift.

**Supplemental Figure 4.**
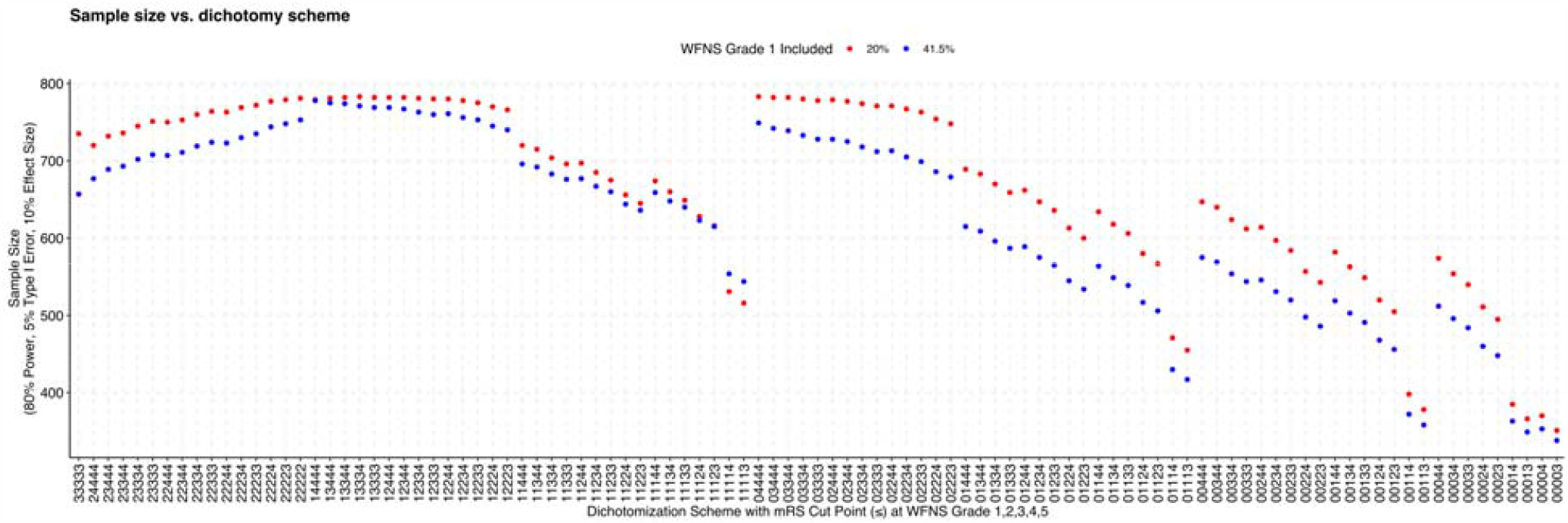
Sample size estimates of 101 dichotomization schemes. The notation, for example, 11234 designates the dichotomizing mRS score (≤) for each WFNS grade sequentially: mRS ≤1 for grade 1, mRS ≤1 for grade 2, mRS ≤2 for grade 3, mRS ≤3 for grade 4, and mRS ≤4 for grade 5. There are two fixed dichotomy schemes: 22222, which equates to mRS ≤2, and 33333, which equates to mRS ≥4 (which is the same as mRS ≤3). Red dots are sample sizes when WFNS grade 1 patient enrollment is limited to 20% from what is expected (41.5%).

**Supplemental Figure 5.**
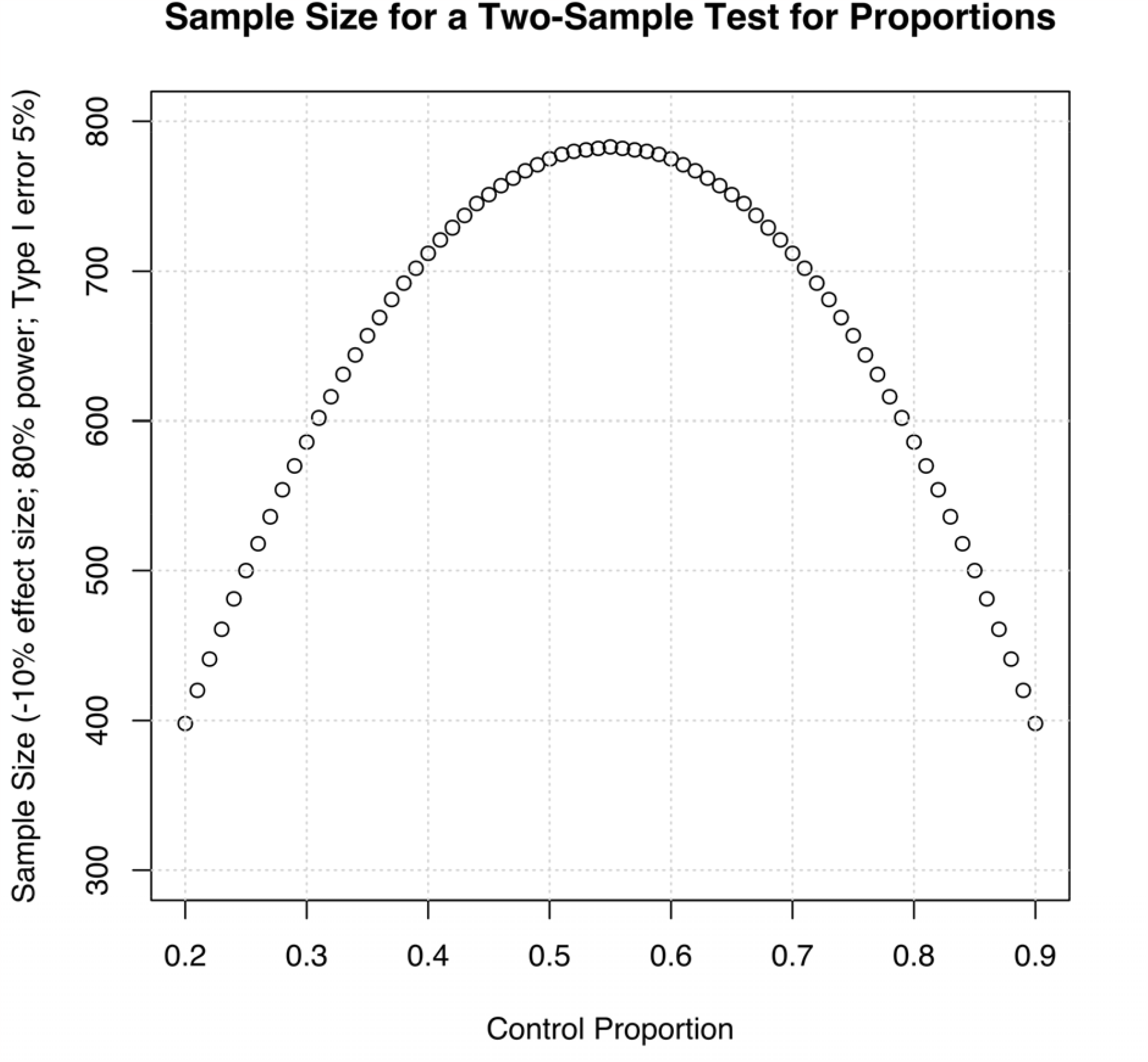
Sample size estimates for a two-sample test for proportions.

## Notes

### Competing Interest Statement

The authors have declared no competing interest.

### Funding Statement

This study was funded by NIH/NINDS U01NS087748 (J.E. and J.B.)

